# Questioning dual-task assumptions in Parkinson’s disease: heterogeneity, cognitive contributions and adaptative changes

**DOI:** 10.1101/2025.10.17.25338216

**Authors:** Malin Freidle, Franziska Albrecht, Hanna Johansson, Erika Franzén

## Abstract

Dual-task impairment is commonly assumed to characterize and challenge people with Parkinson’s disease (PD). We investigated dual-task performance in people with mild to moderate PD (n = 93) and healthy controls (HC, n = 40) using two dual-task paradigms. Contrary to hypotheses, the PD group did not show slower gait or response times more than HC group. In the PD group, longer response times in one task correlated strongly with both accuracy and executive function, suggesting that slower responses may sometimes reflect beneficial adaptations linked to higher cognitive ability. We also observed substantial within-group variability and overlaps between PD and HC across multiple performance outcomes. Exploratory analyses indicated that these unexpected findings largely reflect the influence of cognitive ability on dual-task performance. We caution against assuming changes during dual-tasking are negative and highlight the potential of individualized assessment of cognitive and dual-task abilities for improving clinical practice.

## Introduction

Performing a task in parallel to another, i.e., dual-tasking, is often more difficult for both young and old individuals than to perform the tasks separate from each other ^1–4^. It is often assumed that dual-task impairment is greater for individuals with Parkinson’s disease (PD) and many exercise and rehabilitation programs also stress the importance of enhancing dual-task ability (for an overview see Johansson ^5^). Explanations to why dual-task impairment could be particularly pronounced in people with PD include lowered automaticity in motor skills as well as common impairments in attention and executive functions ^10,11^. A more brain focused explanation claims that the dopamine depletion in people with PD causes dysfunctional overlaps between fronto-striatal circuits and thereby deficits in dual-task performance ^7^. However, empirical studies have not always supported impaired dual-task performance in people with PD compared to healthy participants of the same age and when dual-task differences are found, the outcomes impaired often differ between studies ^6–9^. For now, there is no solid comprehension on if, when and how people with PD are impaired in dual-tasking compared to healthy individuals of a similar age. This is unfortunate as our daily life is full of dual-task situations e.g., walking and talking to a friend, driving a car while finding the correct way or getting dressed while responding to the ringing phone. It is important to gain firm knowledge of whether dual-task abilities are particularly difficult for people with PD, if some individuals with PD have more pronounced problems and what could explain such individual differences. It is also crucial to understand which changes in behaviour during dual-tasking that are problematic and additionally, if some changes might instead help maintain overall performance. Increased knowledge of these aspects has the potential to optimize rehabilitation and intervention development for people with PD.

Our research group has previously found dual-task effects in people with PD in a motor-cognitive dual-task (straight walking in parallel to performing an auditory Stroop, from now on “the Walking task”). We also saw that performance on the Walking task differed between individuals with and without Mild Cognitive Impairment (MCI) ^12^. We will now expand on this finding by comparing dual-task performance in people with PD to healthy adults. Comparisons will be primarily made for the Walking task as well as for a screen-based motor-cognitive dual-task performed during functional Magnetic Resonance Imaging (fMRI) acquisition (from now on “the Scanner task”), see Fig. 1. We will also analyse group differences in the Timed-Up-and-Go task (TUG) as well as group differences in brain activity as measured with fMRI during the Scanner task. Additionally, we will expand on our previous findings on the role of cognitive function for dual-task performance by exploring the relation between executive function and dual-task performance.

**Fig. 1.**
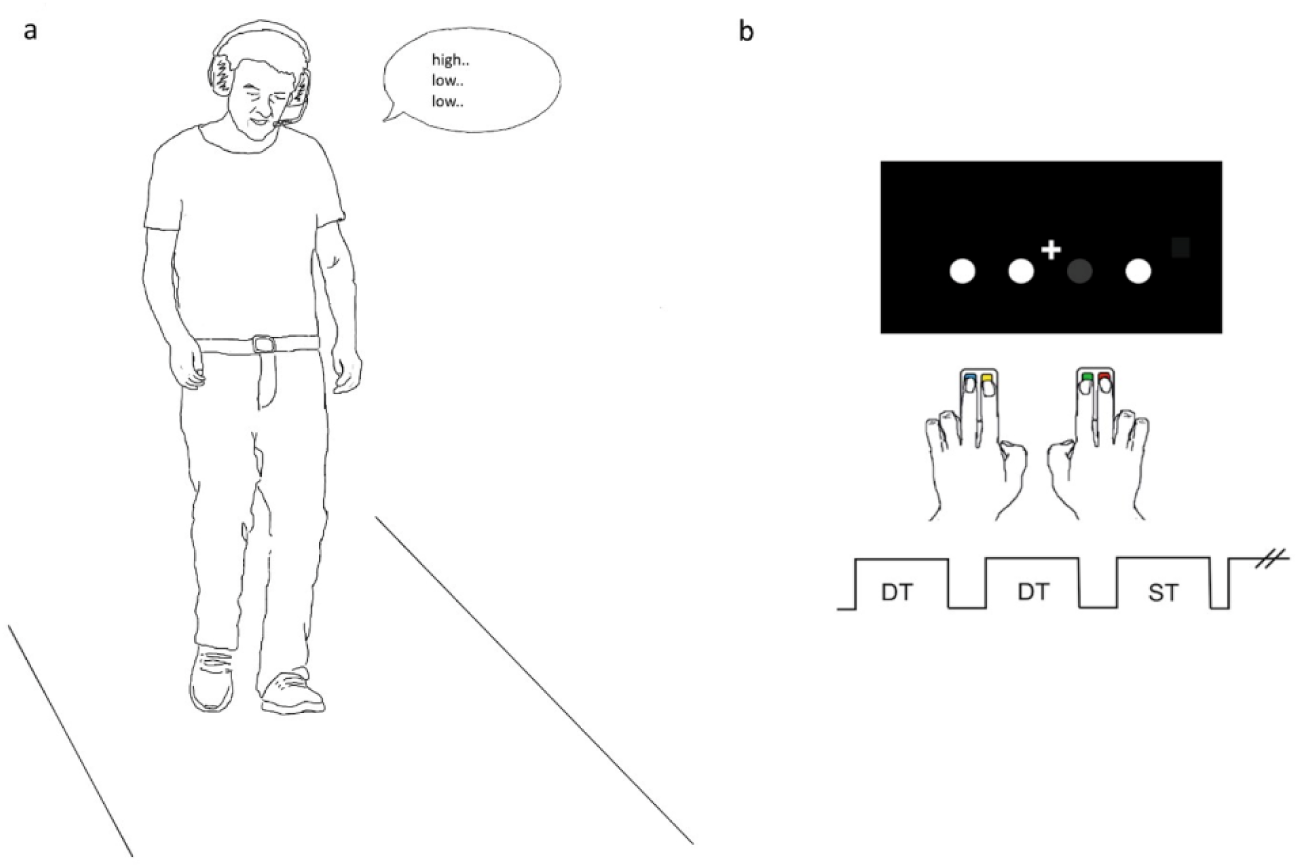
The Walking task and the Scanner task. **a** The Walking task. Participants walked on an electronic walkway system in parallel to hearing and responding to auditory Stroop stimuli through the headset. Both the walking and the auditory Stroop were also performed as single tasks for comparison. **b** The Scanner task. Participants performed a Multiple-Choice task in parallel to counting the plus signs intermittently flashed above the circles. The Multiple-Choice task was also performed block-wise as a single task, marked here as ST. DT = dual-task block.

For the Walking task, we will test the hypotheses that the PD group will show larger dual-task impairments than the healthy control group (HC) for the motor task outcomes gait speed and step time variability and for the cognitive task outcomes reaction time (RT) and RT variability. For the Scanner task, we will test the hypotheses that the PD group will show larger dual-task effects than the HC group for the motor task outcome RT and the cognitive task outcome counting. We will also test whether our fMRI data shows similar results as reported by Nieuwhof et al.,(2017) i.e., more ventro-posterior activity in the putamen during dual-tasking for the PD group than the HC group as well as a negative correlation between ventro-posterior activity in the putamen and dual-task performance in the HC group.

## Results

### Participant characteristics

All participants were ≥ 60 years old and part of the EXercise in PArkinson’s disease and Neuroplasticity trial (EXPANd) ^13,14^. The present analyses are based on baseline data from the EXPANd trial. For the Walking task analyses, 93 individuals with mild to moderate idiopathic PD and 40 healthy individuals were included. Eighty-six participants with PD and 38 HC participants performed the Scanner task used in the MR scanner. Seventy-three participants with PD and 38 healthy individuals were included in the analysis of fMRI data (the missing fMRI data were due to inability to go through with the fMRI, technical issues and excessive movement). To preserve statistical power, all participants who completed the Scanner task were included in the behavioural analyses. Participants remained on their regular dopaminergic medication schedule throughout the study. For more participant information see Table 1 and the Methods section.

**Table 1.** Participants Characteristics.

| Variable | PD (n = 93) | HC (n = 40) |
| --- | --- | --- |
| Sex, female n (%) | 34 (36.6) | 29 (72.5) |
| Age (years) | 71.0 (6.1) | 70.4 (4.9) |
| Body mass index (kg/m<sup>2</sup>) | 25.3 (3.5) | 24.7 (3.1) |
| Education (years) | 14.8 (3.0) | 15.8 (3.9) |
| Montreal Cognitive Assessment (0-30) | 25.8 (2.4) | 26.7 (2) |
| Mini Balance Evaluation Systems Test (0-28) | 20.9 (3.5) | 23.6 (2.5) |
| Levodopa equivalent dose (mg/day) | 554.1<br>(331.3) | n.a. |
| Years with diagnosis | 5.2 (4.5) | n.a. |
| Hoehn & Yahr |  |  |
| 2, n (%) | 71 (76.3) | n.a. |
| 3, n (%) | 22 (23.7) | n.a. |
| MDS-UPDRS, Part III, motor examination (0-132) | 31.1 (11.2) |  |

MDS-UPDRS: Movement Disorder Society-Unified Parkinson's Disease Rating Scale.
n.a. = not applicable. PD = Parkinson's disease. HC = healthy controls.

### The Walking task

For the primary outcome gait speed, the task type (single = 0/dual = 1) by group (PD = 1/HC = 0) interaction effect was estimated to b= 0.04 (−0.01, 0.09)), p= 0.13 (Table 2). Descriptively both the PD and the HC group walked slower during dual-tasking (than during single-tasking) with a DTE (dual-task effect) group difference of 3 percent points in the direction that the HC group slowed down more than the PD group. There were however differences within the PD group as well as overlaps with the HC group. The correlation between executive function and DTE gait speed was estimated to rho = 0.42 (0.23, 0.58) i.e., higher executive functions were associated with slowing down less during dual-task performance in the PD group (Fig. 2). Relatedly, for the PD group there was a negative correlation of −0.25 (−0.44, −0.04) between DTE gait speed and the Stroop RT during the dual-task and in the HC group the correlation was estimated to −0.44 (−0.66, −0.14). These negative correlations indicate that more decrease in gait speed is associated to slower response to the cognitive task.

**Fig. 2.**
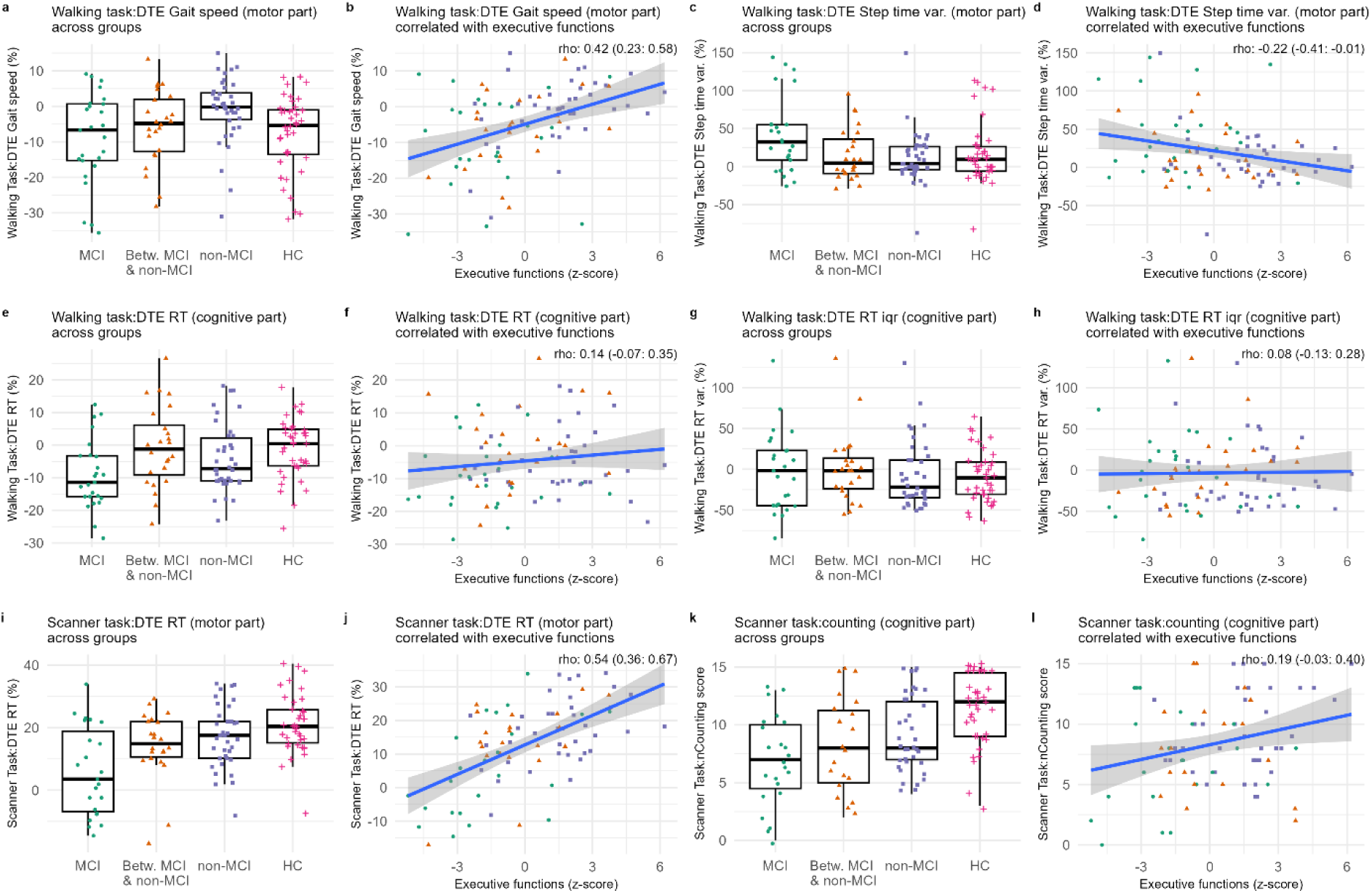
Dual-task performance across groups and in relation to executive function. For all boxplots, the PD group is presented classified as with MCI (Mild Cognitive Impairment) or without (non-MCI), or in between those groups (Betw. MCI & non-MCI). HC = healthy control group. All correlation plots are within the PD group. DTE = dual-task effect. rho = Spearman’s rang order correlation coefficient. Var. = variability, RT = reaction time, corr. = correlation. **a-h** Primary outcome variables of the Walking task. DTE Gait speed: A negative value indicates lower speed during the dual-task than the single task. DTE Step time var: a positive value indicates more variability during dual-task. DTE RT = A positive value indicates slower responses during dual-task. DTE RT var. = A negative value indicates more variability during dual-task. Executive functions (z-score) = a higher value indicates higher executive functions, **i-l** Primary outcome variables of the Scanner task. DTE RT = A positive value indicates slower responses during dual-task. Counting = a positive value indicates more correct responses. Executive functions (z-score) = a higher value indicates higher executive functions Counting was only performed during the dual-task.

**Table 2.**
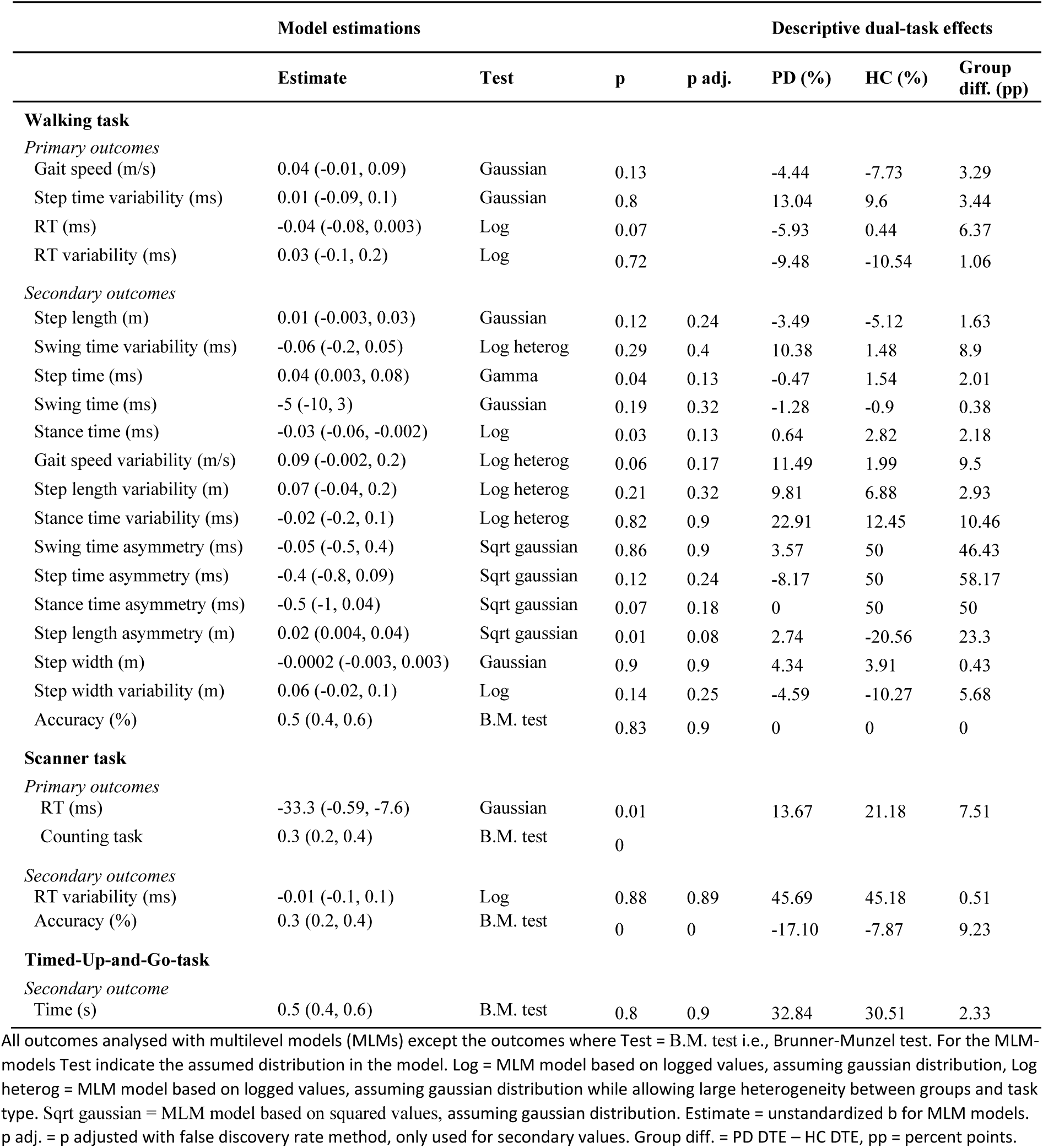
Dual-task outcomes.

However, a funnel shape of the data points requires cautious interpretation of these correlations (Fig. 3). The median of the accuracy outcome was 97% for the PD group and 100% for the HC group. Because of this strong ceiling effect, a correlation coefficient is not a good representation of the relation between DTE gait speed and Stroop accuracy.

**Fig. 3.**
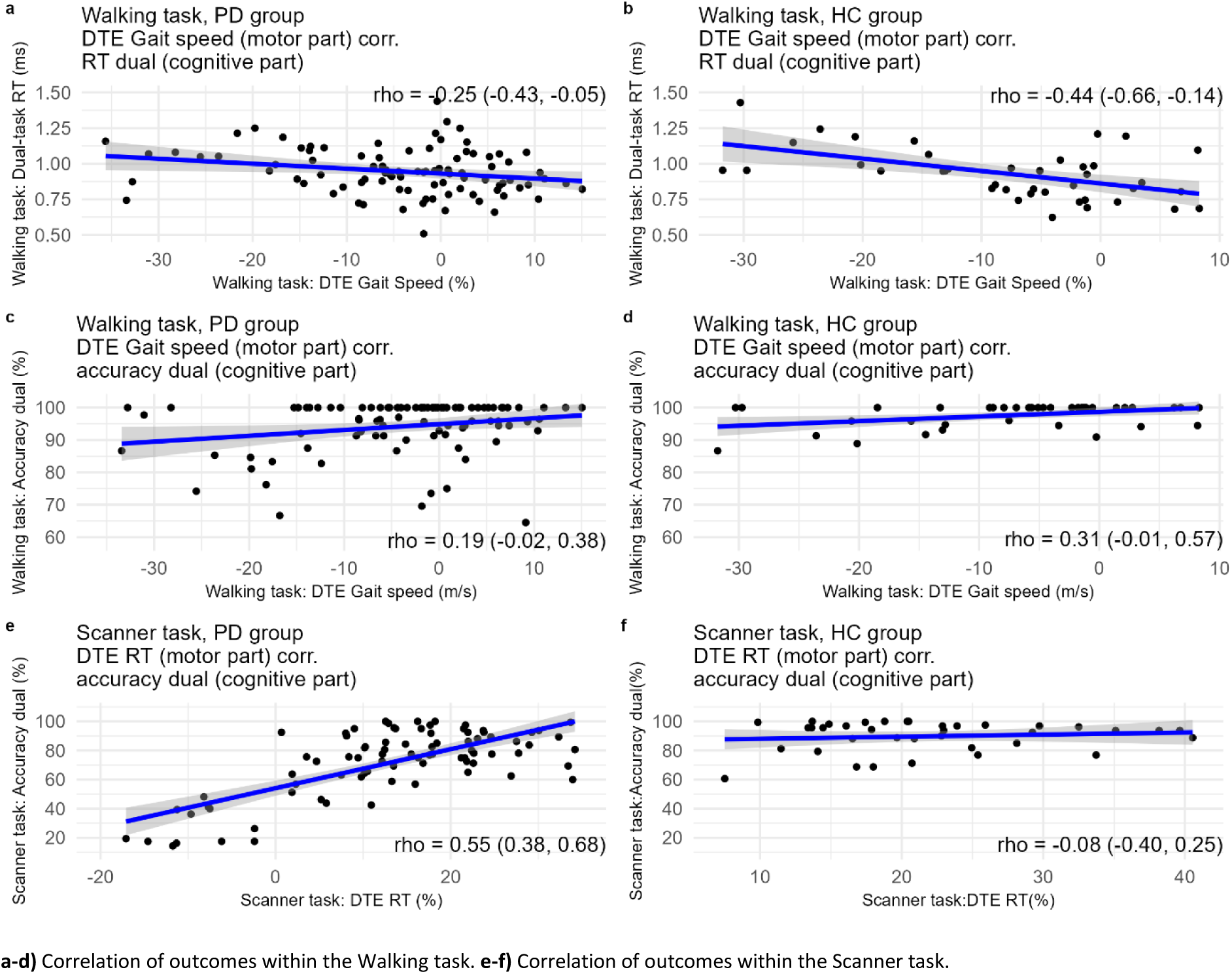
Correlations between gait speed, RT and accuracy outcomes. **a-d)** Correlation of outcomes within the Walking task. **e-f)** Correlation of outcomes within the Scanner task.

For the primary outcome step time variability, the task type by group interaction term was estimated to b = 0.01 (−0.09, 0.1), p = 0.8. Descriptively both the PD and HC group increased their step time variability during dual-tasking with a DTE group difference of 3 percent points in the direction that the PD group increased step time variability more than the HC group. There were however differences within the PD group as well overlaps with the HC group. The correlation between executive function and DTE step time variability estimated to rho = −0.22 (−0.41, −0.01) i.e., higher executive functions were associated with a smaller increase in step time variability during dual-task performance in the PD group.

For the primary outcome Stroop RT, the task type by group interaction effect was estimated to b = −0.04 (−0.08, 0.003), p = 0.07. Descriptively, the PD group decreased their RT during dual-tasking and the HC group had a similar RT during single and dual-task with a DTE group difference of 6 percent points but with group overlaps. The correlation between executive function and Stroop DTE RT in the PD group was positive and estimated to 0.14 but the CI included 0 (−0.07, 0.35).

For the primary outcome Stroop RT variability, the task type by group interaction effect was estimated to b = 0.03 (−0.1, 0.2), p < 0.72. Descriptively, both groups decreased their RT variability during dual-tasking and the DTE group difference was small (1 percent point in the direction that the HC group decreased RT variability more) and with group overlaps. The correlation between executive function and Stroop DTE variability in RT in the PD group was estimated to 0.08 but the CI included 0 (−0.13, 0.28). See Fig. 2 for visualisations of the primary Walking task outcomes.

The secondary outcomes of the Walking task included Stroop accuracy and another 14 gait variables. All analyses of group differences for these secondary outcomes were non-significant after using the false discovery rate correction method for multiple testing. The DTE difference between the PD and HC group was > 10 percent points for stance time variability and all four asymmetry outcomes. There were negative correlations > −0.3 (CIs not including 0) between executive functions and the secondary Walking DTE outcomes swing time, swing time variability, step time, and stance time i.e., higher executive functions were associated with smaller increases in these gait variables during dual-tasking. There was a positive correlation > 0.3 (CI not including 0) between executive functions and DTE step length i.e., higher executive functions were associated with less increase in step length during dual-tasking.

The complementary multilevel models (MLMs) within the PD group showed similar effects of executive function as the correlations between executive functions and DTE outcomes (Supplementary Fig. 2). Outcomes for all Walking task models and the descriptive DTE effects are presented in Table 2. See Supplementary Table 1 and 2 for descriptive data for all single and dual task versions and MLM analyses not excluding extreme values as well as Supplementary Fig. 1 for plots of secondary Walking task outcomes.

#### Prioritization patterns and their effects

The difference in prioritization pattern reported by Johansson^12^ was overall replicated based on the our refined analyses of the auditory Stroop i.e., the PD MCI group descriptively showed more cognitive prioritization (m=16.5, sd = 14.7) than the PD non-MCI group (m= 2.1, sd = 11.2). We also observed an overall cognitive prioritization (m=7.2, sd = 13.2) in the HC group. We found no significant and/or larger correlations between Walking DTE RT and outcomes often assumed to represent gait instability including step width variability and other variability and asymmetry outcomes (Supplementary Fig. 3).

### The Scanner task

For the primary outcome RT, the task type by group interaction effect was estimated to 0.09 (0.06, 0.1), p < 0.00. Descriptively, both the PD group and the HC group overall increased RT during dual-tasking but with a group DTE difference of 7.5 percent points in the direction that the HC group increased RT more than the PD group. There were however differences within the PD group and overlaps with the HC group. The correlation between executive function and DTE RT was estimated to 0.61 (0.45, 0.73) i.e., higher executive functions were associated with a larger increase in RT during dual-task performance in the PD group (Fig. 2). Relatedly, the range in accuracy for the PD group was 14-100% (median = 77%) and there was a positive correlation between DTE RT and the accuracy during the dual-task of 0.54 (0.36, 0.67) meaning that a higher increase in RT during dual-tasking was associated with higher accuracy scores in the PD group. The accuracy range in the HC group was 37-100% (median = 94%) and the correlation between DTE RT and accuracy was estimated to −0.08 (−0.4, 0.25) i.e., no association between the variables but generally high accuracy scores (Fig. 3).

For the primary outcome scores on the counting task, the Brunner-Munzel test group difference estimate of the DTE was 0.3 (0.2, 0.4), p < 0.00. The HC group had a median score of 12 (interquartile range (iqr) = 5.5) and the PD group a median score of 8 (iqr = 5.25; there was no DTE outcome of the counting task as it was only present during the dual part). The correlation between executive function and the scores on the counting task was positive and estimated to 0.19 but the CI included 0 (−0.03, 0.40).

For the secondary outcome RT variability, the task type by group interaction effect was estimated to −0.01 (−0.1, 0.1). Both the PD and HC group increased RT variability during dual-tasking with a minimal DTE group difference. There were however differences within the PD group and an overlap with the HC group. The correlation between executive function and DTE RT variability estimated to 0.37 (0.17, 0.55) i.e., higher executive functions were associated with a larger increase in RT variability during dual-task performance.

For the secondary outcome accuracy, the Brunner-Munzel test group difference estimate of DTE was 0.7 (0.6-0.8). Both the PD and the HC group decreased their accuracy during dual-tasking with a DTE group difference of 9.0 percent points in the direction that the PD group decreased their accuracy more than the HC group during dual-tasking. The correlation between executive function and DTE accuracy was positive and estimated to 0.19 but the CI included 0 (−0.03, 0.39). Complementary MLM analyses within the PD group yielded effects of executive function comparable to those from the correlation analyses (Supplementary Fig. 2).

### The TUG task

For TUG, the Brunner-Munzel test group difference estimate of DTE was 0.47 (0.38-0.6). Both the PD and the HC group increased their time walking during dual-tasking with a DTE group difference of 2.0 percent points in the direction that the PD group decreased their accuracy more than the HC group during dual-tasking.

### Task comparisons

The Spearman correlations between the DTE outcomes of the three tasks were all rho < 0.15, see Fig. 4. Correlations between single and dual outcomes between the three tasks e.g., Scanner single task RT and Walking single task gait speed, were generally higher (e.g., −0.64 for dual-task gait speed and motor TUG). More correlations can be found in Supplementary Table 3.

**Fig. 4.**
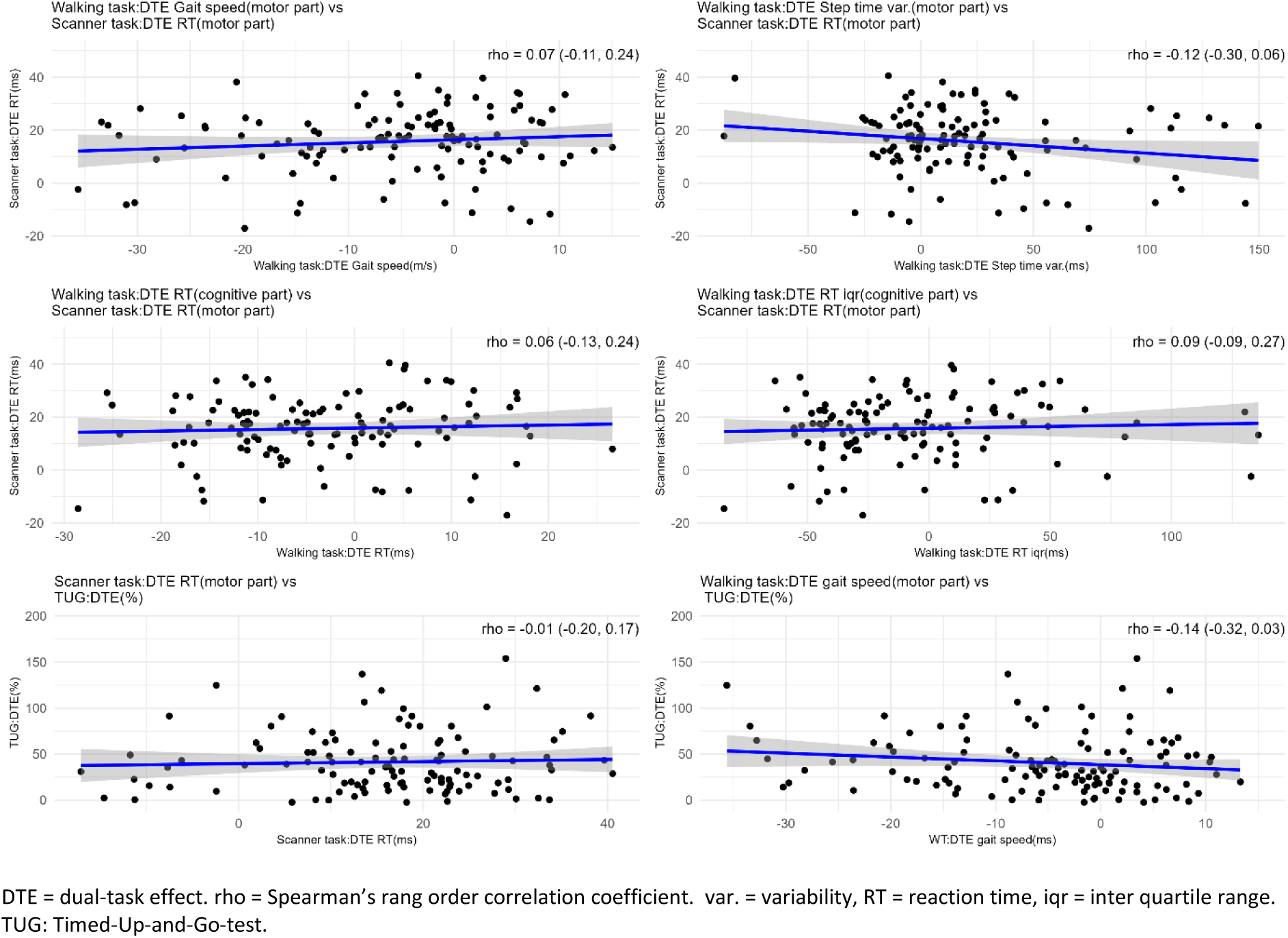
Relations between the task DTE outcomes. DTE = dual-task effect. rho = Spearman’s rang order correlation coefficient. var. = variability, RT = reaction time, iqr = inter quartile range. TUG: Timed-Up-and-Go-test.

### fMRI data

The PD group did not have statistically significantly higher ventro-posterior activity in the putamen than the HC group (no significant clusters of voxels). Additionally, the PD group did not have statistically significantly higher activity in any of the remaining three parts of the putamen compared to the HC group. Nor was there a significant or large correlation between activity in the ventro-posterior putamen and performance on the Scanner task within the HC group, rho estimated to 0.22 (−0.11, 0.52), p = 0.19 (Supplementary Fig. 4).

## Discussion

We investigated dual-task performance in people with mild to moderate PD and in older healthy adults, focusing on a Walking task and a screen-based task performed in an MR scanner. Our hypotheses that the PD group would slow down significantly more than the HC group when switching from a single task version to the dual-task version for both the Walking task (gait speed and RT to auditory stimuli) and the Scanner task (RT to visual stimuli), were not supported. Instead, data indicated that the PD group slowed down less than the HC group, especially in the RT outcomes of both tasks. For the PD group, increased RT in the Scanner task showed strong positive correlations to both accuracy and executive functions, suggesting that longer response times can sometimes be a beneficial adaptation done by those with higher cognitive ability. Our other primary hypotheses were not supported either - with the exception that the PD group performed worse than the HC group on the counting part of the Scanner task. Importantly, there were large ingroup variances as well as non-negligible overlaps between the PD and HC group for multiple outcomes. Exploratory analyses indicate that cognitive function show important relations to dual-task performance in both expected and unexpected ways.

Dual-task impairment is commonly assumed to be greater for people with PD and to constitute a large part of observed motor problems ^15–17^. Relatedly, multiple rehabilitation programs for people with PD focus on enhancing dual-task ability ^5^. However, not to be forgotten, dual-tasking has performance effects also for healthy older and younger adults ^3,4,18^. When comparing dual-task performance in PD to healthy older individuals, some studies show PD specific deficits while others do not ^6–9^. This inconsistency is partly in line with our results where a few outcomes show statistically significant more dual-task impairment for the PD group, a few analyses show close to significant group differences but the majority of analyses are non-significant and do not suggest large differences between the PD and HC group. In addition to discussing our results, we will provide plausible explanations for the inconsistency in previous studies.

Earlier work by our research group showed that dual-task performance in PD varies by cognitive ability as measured by the dichotomous classification into MCI and non-MCI ^12^. In the present study we extended this finding by 1) presenting results for an additional dual task in the Scanner, 2) including healthy older individuals and 3) relating the dual-task performance in the PD group to executive functions specifically. We observed some group level differences but also non-negligible overlaps between the PD and HC group i.e., for all outcomes there were healthy participants who performed at the same level as PD participants, even PD participants categorized as having MCI (see Boxplots in Fig. 2). There were also moderate to strong correlations between executive functions and multiple of the DTE outcomes for the PD group. Taken together, the data strongly indicate that an individual with a PD diagnosis should not per se be assumed to present with dual-task effects greater than a healthy older individual. Rather than a PD diagnosis, cognitive function may serve as a more sensitive indicator of dual-task impairment for an individual. We did however only include people with mild to moderate PD and it is reasonable to assume that differences in dual-task performance to a HC group are more pronounced in more severe PD where cognition is also more often severely impacted ^19^.

Our results also call for a discussion on what behavioural changes during dual-tasking should be expected and/or interpreted as problematic. In contrast to our hypotheses for the Walking task, we did not observe a larger decrease of gait speed in the PD group than the HC group. Even more unexpectedly, on group level, participants with PD actually decreased their RT i.e., responded faster, when performing the auditory Stroop in parallel to walking (in contrast to sitting down) while the HC group showed a very small difference between single and dual-tasking RT. Both the PD and HC group also unexpectedly decreased their RT variability when dual-tasking. Because RT in the Walking task is the outcome for the cognitive task, decreased RT and RT variability during dual-tasking could be an indicator of cognitive task prioritization, often interpreted as an increased risk for falls. In our reanalysis of prioritization patterns as reported in Johansson et al., ^12^ we found that especially PD with MCI, and the HC group, tended to prioritize the cognitive task over the gait task, often interpreted as an increased risk for falls. However, there were only small, non-significant correlations between Walking task DTE RT (cognitive outcome) and commonly assumed indicators of gait instability, such as variability or asymmetry ^20,21^. Relatedly, no strong correlations were found between executive function and Walking DTE RT or DTE RT variability in the PD group, and about half of the HC participants also showed reduced RT and RT variability during dual-tasking. In total, it is far from clear that decreased RT or RT variability has negative effects on gait performance.

In the Scanner task, we saw a similar but intensified unexpected RT pattern such that the PD group increased their RT less than the HC group during dual-tasking. There was also a strong correlation between increased RT during dual-tasking and accuracy in the PD group. This raises the possibility that for some tasks slowing down when an additional task is added might be beneficial for overall performance i.e., in the case of the Scanner task, that slowing down during dual-tasking is a prerequisite for maintained accuracy. The need to slow down likely becomes more pronounced as task difficulty increases and individuals approach their cognitive limits. This may explain why RT increased more during dual-tasking in the Scanner task compared to the Walking task, supported by the lower accuracy observed in the Scanner task indicating its greater difficulty. Additionally, the strong positive correlation between executive function in PD and increased RT during the Scanner task suggests that higher cognitive function may be necessary either to recognize when slowing down is needed or to effectively inhibit rapid button presses, thereby maintaining overall performance in more demanding tasks. The positive correlation between executive function and RT DTE variability in the Scanner task is harder to explain. One possibility is that the more successful test taker (with higher executive functions) prolongs pressing the correct button of the visuo-motor part in the Scanner task whenever the symbol to count (additional task) appears, to in that way increase overall performance. When there is instead nothing to count i.e., during the single task, the RT is more stable.

Altogether, our data show that dual-task effects do not consistently appear as expected, with substantial intraindividual variation evident in both PD and HC populations. Our data demonstrate that cognitive abilities play a significant role in dual-task performance in the PD group and while the EXPANd trial did not include cognitive testing for the HC group, previous studies in healthy individuals have similarly demonstrated an impact of cognitive abilities on dual-task performance ^22^ However, other factors likely contribute. Our group has previously reported that individuals with PD differ in whether they are predominantly cognitively or motorically affected, which may account for part of the variance ^23^. In healthy populations, personality traits such as neuroticism, extroversion and conscientiousness influence dual-task performance e.g., individuals higher on the neuroticism trait tend to show larger gait speed reduction during dual-tasking than those low in neuroticism ^22,24^. Personality may similarly affect performance in PD, but this is for future studies to further investigate.

Further empirical work is also needed to determine whether associations between slowing down (and potentially increasing variability) and maintaining overall performance truly reflect causal mechanisms. Such studies could employ A-B-A-B designs ^25^ in which individuals would be alternately instructed to decrease (A) or increase (B) response times or gait speed during dual-tasking, while monitoring their accuracy or gait instability. These could also include varying difficulty to test our hypothesis that increased task demand require greater slowing down to preserve overall performance. All in all, researcher nor clinicians should not assume that someone with PD has marked dual-task impairments, that behavioural changes during dual-tasking are inherently negative or that superficially similar behaviours (e.g., slowing down) have uniform effects across different dual-tasks and contexts. This highlights the possibility for greater rehabilitation benefit effects if interventions are more tailored to the individual, informed by in-depth assessments of cognitive ability and dual-task performance under varying instructions and contexts. For example, for some individuals and in certain situations, it may be advantageous to explicitly encourage slowing of gait speed or response times during dual-tasking to enhance safety and/or overall performance.

The inconsistency in the literature on dual-task impairments in PD may partly be explained by our findings. Differences in cognition and disease progression within the PD population likely produce divergent outcomes. For example, studies including more people with PD with advanced cognitive problems are more likely to detect differences from healthy individuals than those with predominantly cognitively preserved participants. Moreover, the somewhat contrasting results between our two dual-tasks suggest that task type and difficulty can elicit different behavioural adaptations—meaning what is deemed an impairment in one task should not be so in another. These variations across studies, combined with the small and often underpowered samples common in PD dual-task research ^7,8^, likely contribute to the literature inconsistencies. Our data further show low correlations between DTE outcomes from three different tasks (Walking task, Scanner task, TUG), which could imply they measure distinct abilities, though such strong segregation is theoretically difficult to justify. A more plausible explanation is the reduced reliability of DTE difference scores— known to attenuate observed correlations—compared to the reliability of the original single- and dual-task outcomes, as supported by prior empirical work ^26^ ^27^. Studies primarily using the more unreliable DTE outcomes plausibly loose statistical power, and thereby produce less robust results, contributing to the literature inconsistency. This recognition of DTE reliability limitations guided our choice to use MLM analyses and avoid difference scores in primary analyses while keeping them for pedagogical illustrations of outcomes across groups and in correlation to executive function.

As for the fMRI analyses, we found no significant group differences in brain activity, nor support for the results by Nieuwhof ^7^ regarding greater ventro-posterior putamen activity during dual-tasking in the PD group compared to the HC group. These null findings may well reflect a true lack of group differences. While our statistical power was uncertain and task fMRI studies are known to require large samples ^28^, it is still possible that some effects went undetected. However, since Nieuwhof ^7^ employed an even smaller sample size, and differences in study design/analyses and pre-processing exist, it is understandable that their findings have proven difficult to replicate.

Study limitations include the above discussed power of the fMRI analyses, the ceiling effect in the Walking task accuracy outcome and the lack of extensive cognitive testing for the HC group. Analyses based on variance in Walking task Stroop accuracy as well as measures of executive functions in HC, could further have illuminated the complicated patterns found. Relatedly, for clear visualization we correlated executive function with the DTE outcomes even though DTE outcomes are generally less reliable, causing attenuated correlations. However, our complementing MLM analyses on the effect of executive function, avoiding the problems with DTE outcomes, showed similar results as the DTE correlations. Strengths of the study include an unusually large sample including both participants with PD as well as healthy participants of a similar age, preregistration of primary analyses, analyses of multiple motor and cognitive outcomes for three different dual-tasks as well as illuminating complementary analyses and visualizations.

Altogether, our findings provide a comprehensive overview of dual-task ability in people with PD compared with healthy older controls. For most outcomes −including all our primary outcomes-there were no significant or substantial impairments in the PD group relative to the HC group. Contrary to our hypotheses, the PD group slowed down less than the HC group, particularly in RT outcomes for both the Walking and the Scanner task. Notably, we demonstrated substantial within-group variability and considerable overlaps between the PD and HC group for multiple outcomes. Exploratory analyses suggest that these unexpected results are largely attributable to the influence of cognitive function on dual-task performance. In light of this, we caution against assuming expected behavioural changes in people with PD during dual-tasking or automatically judging them as detrimental to overall performance. Our findings have important implications for both research and clinical practice: rehabilitation may achieve greater benefits when guided by in-depth, individualised assessments of cognitive ability as well as dual-task performance across different tasks and contexts.

## MATERIALS AND METHODS

### Participants

All participants with PD had a documented diagnosis of idiopathic PD as assessed by their neurologist. The EXPANd project (registered at ClinicalTrials.gov: NCT03213873, 2017-07) consisted of a randomized controlled trial (RCT) with pre- and post-assessments for people with PD as well as assessments of a HC group. In the present study, only the pre-assessment data for the participants with PD was used and the HC participants were assessed parallel to the pre-assessments of the RCT. This means that there were no differences in the exposure to either the situation or the task between the participants with PD and the HC group for the data used for the present analyses.

Inclusion criteria in the EXPANd trial for the individuals with PD were: mild to moderate PD severity defined as Stage 2 or 3 on the Hoehn & Yahr scale ^29^, age ≥60 years, a score ≥21 on the Montreal Cognitive Assessment (MoCA) ^30^, and no other disease or symptom that could significantly affect balance or gait. More details on the inclusion/exclusion criteria for individuals with PD can be found in Franzén et al.^14^. Inclusion criteria for the HC participants were age ≥60 years, a MoCA score ≥23, and no disease or symptom that could significantly affect balance or gait. The lower MoCA cut-off of ≥21 for the participants with PD was chosen to recruit a representative sample of the PD population where cognitive deficits are common in people with PD already in the mild to moderate stages ^31^.

### Procedure and task design

Participants with PD were tested in the ON state to reflect daily-life context. Assessments of gait, cognitive ability and the MR scanning occasion were conducted on separate days within a week to avoid fatigue.

During the Walking task, the participants conducted a gait task and an auditory Stroop task separately (single task) and in combination (dual task). Gait was assessed using an electronic walkway (GAITRite, active zone: 8.3 m, CIR Systems, Inc), with participants walking at a self-selected pace back and forth eight times per condition; the first two trials served as practice. All began with the single gait task. The auditory Stroop presented the Swedish words for ‘high’ and ‘low’ in congruent or incongruent tones via wireless headphones, and participants responded verbally as fast as possible to the tone pitch regardless of the word. They completed two practice trials (or more if needed for task comprehension) of the Stroop seated, then two while walking. Participants were randomized to begin with the auditory Stroop as either a single or dual-task. They were instructed to pay equal attention to both tasks when performing them in parallel. For more details see Johansson et al. ^12^.

During the MR scanning, the participants performed a Multiple-Choice RT task without (single task) and together with a simple counting task (dual-task). This task consisted of eight blocks á 32 trials: three single blocks with only the Multiple-Choice RT task and five blocks with the additional counting task (order of blocks; S1 – D1 – D2 - S2 – D3 - D4 - D5 – S3). Each block was followed by a six-second-long break. For all blocks, four white circles on a horizontal line were shown on a black screen and each circle’s position corresponded to one of the four buttons of the two response pads that were placed on the participant’s lap. Every 1.2 s, one of four circles turned grey, prompting participants to press the corresponding button as quickly as possible. Participants used the index and middle fingers of both hands to respond. In dual blocks, a plus sign appeared above the circles five to ten times per block, and participants counted these while performing the RT task. After each dual block, they had six seconds to select the correct number of plus signs from four alternatives (ranging from 0 to 7 integers from the correct alternative). Each dual block began with all circles briefly turning green to signal its start and avoid surprise and accompanying movement. As in the Walking task, participants were instructed to give equal attention to both tasks.

The participants practiced the Scanner task before they entered the scanner. The practice ended when the participant achieved 80% accuracy or after a maximum of three blocks consisting of 32 trials each. The task was presented in PsychoPy (version 1.85.4). During the fMRI acquisition, the task was presented using a computer screen positioned behind the scanner. A mirror inside the scanner head coil enabled the participants to view the task. The total time inside the scanner was approximately 40min. The dual-task was 7min long and presented as the third MR sequence.

During the gait assessment occasion, participants also performed the TUG test that assesses functional mobility. It consisted of assessing the time it took for the participant to stand up from a chair, walk 3 meters, turn around, walk back to the chair, and sit down. TUG was only included as a secondary study outcome.

On a separate day from gait testing as well as the scanner task, the participants with PD underwent an extensive cognitive test battery that form the basis for our measure of executive function. As the cognitive function of the HC group was not the focus of the EXPANd project, only the PD group completed the cognitive test battery.

The HC participants received two cinema vouchers after participation while the participants with PD participated as part of the RCT where they attended interventions aimed to ameliorate PD-related symptoms. After providing oral and written study information, written consent was obtained from all participants. The study was approved by the Regional Research Ethics Board of Stockholm (2016/1264–31/4 with amendments).

### Acquisition of the fMRI data

Indirect measures of brain activity were acquired by fMRI and the blood-oxygen-level-dependent (BOLD) signal. A 3T Phillips Ingenia scanner with a 15-channel head coil with the following parameters was used: repetition/echo time (TR/TE) =2085/35ms, flip angle=75°, voxel-size: 3.5×3.5×3.5mm, the field of view: 224×224×140, 265 slices in ascending order.

### Statistical analyses

For all outcomes, all values 3 standard deviations (sd) over or under the mean were replaced by NA to avoid biased estimates. All MLM-analyses were additionally run with all values included (see supplement).

#### Analyses of the Walking task

Our preregistered primary outcomes for the Walking task were gait speed, step time variability, response times and variability in response times to the auditory Stroop. For both the primary and secondary analyses, we used preregistered MLMs with task type (single vs. dual), group (PD vs. HC), and their interaction as fixed effects, and a random intercept. The distribution for each model (e.g., Gaussian) was chosen based on residual diagnostics. No MLM met residual assumptions for the Walking Stroop accuracy outcome. Therefore, we used the Brunner–Munzel test, which does not assume normality or equal variances, to compare DTE accuracy between groups.

For the four primary outcomes of the Walking task, the alpha level was set to 0.05. For the remaining 14 gait variables analysed, we corrected for multiple comparisons by using the false discovery rate method with alpha set to 0.05. The variability outcomes for the gait outcomes were calculated as the mean of the variance of the left and right foot. Gait asymmetry was calculated as the left foot value minus the right foot value. Variability in auditory Stroop response times was defined as the iqr of response times. Participants with <60% accuracy on the auditory Stroop were excluded (n=2) to remove those likely not attempting both tasks in parallel. This exclusion was not applied to the Scanner task, as it would have removed 20 participants with PD, substantially reducing power; given its higher difficulty, the 60% threshold may also be too stringent.

In addition to the MLM analyses, we summarised DTEs for the PD and HC groups. DTEs were calculated as ((dual task value - single task value) / single task value) × 100. Outcomes with approximately normal distributions were reported as mean ± SD; others as median and iqr. Boxplots were created for each DTE outcome, grouped by HC and PD subdivided into PD with MCI, PD without MCI, and PD classified as intermediate between non-MCI and MCI. For details on the MCI classification see Johansson et al., ^12^. To further examine cognitive influences on dual-task performance in PD, we correlated a composite executive function score with all DTE outcomes. Because DTE values have limited reliability, for our main outcomes we also ran MLMs within the PD group, including task type (single/dual), executive function, and their interaction as predictors. The executive function measure complements the MCI-based classification by providing a continuous variable—offering greater statistical power and specifically targeting a cognitive domain frequently impaired in PD ^32^. The composite measure of executive function was created using confirmatory factor analysis including four cognitive tests from the Delis–Kaplan Executive Function System and the Wechsler Adult Intelligence Scale. To enable this composite measure, all test values were standardised i.e., converted to a z-score. A positive z-score on the composite measure indicates higher executive functions. for more details see Freidle et al.^13^.

During preparation of the present article, a methodological issue in the Walking task RT analysis for the PD group in Johansson^12^ was detected and so we reanalysed the RT in the Walking task as well as the analysis of prioritization of motor and cognitive outcomes. In addition, we examined correlations between Walking task DTE RT and gait variability and asymmetry, two gait characteristics previously linked to gait instability^20,21^. Strong correlations here would support the idea that allocating greater attention to the cognitive task may come at the expense of gait stability.

#### Analyses of the Scanner task

MLMs with group (PD/HC) and task type (single/dual) and their interactions was used to analyse RT as well as the variability of RT outcome. Due to the non-normal distribution of RT, variability in response times were defined as the iqr of the RT. The MLM models included a random intercept and model distribution was based on residual diagnostics. Non-responses were coded as incorrect responses. Responses with an RT <100 ms were excluded (and coded as missing values) as 100 ms has been reported to be the minimum time for physiological processes such as the perception of stimuli^33^.

No MLM version produced acceptable residuals for the accuracy outcome of the Scanner task and hence a Brunner-Munzel test was used to compare the accuracy DTE outcome for the groups. The counting task was also analysed using a Brunner-Munzel test. DTE values were estimated in the same way as for the Walking task outcomes and the boxplot and correlation plots are presented also for these outcomes.

#### Analyses of the TUG task

No MLM model produced acceptable residuals for the TUG outcome and hence the Brunner-Munzel test, not assuming normal distribution nor equal variances, was used to compare the groups.

#### Task comparisons

Spearman correlations were used to compare the three dual-tasks Walking task, Scanner task and the TUG, to each other. Focus for these comparisons was the DTE outcomes as these are most often used as the primary outcome, but correlations for single and dual-task versions are presented in the supplement.

### Analyses of the fMRI data

Initial quality control of the MRI data was done using MRIQC and the preprocessing was done using fMRIPrep ^34,35^. The first and second-level analyses of the within-region analyses were performed in SPM 12 (version 7765) ^36^. The independent variables for the first-level analyses were the experimental timeline convoluted with the canonical hemodynamic function, 24 motion-derived regressors as well as the first five aCompCor regressors and the cosine regressors.

The masks of our predefined ROIs were based on atlases. To test our hypothesis based on results from Nieuwhof et al,. ^7^ we created four masks of the putamen in the same way. We used the AAL atlas and extracted the areas 77, 78. Then we placed a box halfway between the outer side and the middle of the putamen (between y=0 and y=max length), to separate the putamen into ventral and dorsal posterior as well as ventral and dorsal anterior. Using these masks, we first investigated our primary hypothesis that the PD group should have greater activity in the ventro-posterior putamen than the HC group during dual-tasking. Additionally, we compared the activity for the remaining three parts of the putamen between the HC and PD group. We used independent two-sample t-tests using the contrast images (dual–single) from the first-level analyses to compare the two groups’ task-induced activity. A cluster-defining threshold with a cluster-creating threshold of uncorrected p < 0.001 (voxel-level) and cluster extent threshold of p < 0.05, family-wise error corrected (FWE), was used.

For our secondary hypothesis—that activity in the ventro-posterior putamen would be negatively correlated with dual-task performance in the HC group—we first extracted each participant’s brain activity values. For each HC participant, we calculated a RT difference score between the dual- and single-task blocks of the Scanner task. We then correlated the brain activity values with the RT difference scores using Spearman’s rank-order correlation.

## Data availability

The datasets analysed during the current study are available from the corresponding author on reasonable request.

## Code availability

Analysis plan with amendment, scripts, brain masks and all files needed to run the DT multiple choice task can be downloaded at https://osf.io/fqup9/.

## Deviations from the preregistration

Because the MLM models for the RT outcome of the dual-task did not converge with a random slope, the random slope was excluded from the MLM models.

## Acknowledgments

We would like to thank all our participants for their time. We are grateful to the staff at the MR unit and the Jonasson centre for medical imaging. We would also like to acknowledge Petra Koski for her great work on the logistics of the study including the recruitment, screening and support in assessments. We thank William H. Thompson for help with preprocessing and quality assessing the fMRI data and Lucian Bezuidenhout for preprocessing the Stroop audio files. We thank Elvira Granqvist, Sanna Asp, Mattias Söderberg, Helena Karlfeldt for assisting in data collection and Alexander V. Lebedev and Martin Lövdén for help in developing the Scanner task. We also thank Heather Martin for her invaluable help with the MR safety decisions. ChatGPT and Microsoft co-pilot were used for increasing readability of specific sentences and shorter paragraphs. GitHub Copilot for RStudio has been used for assistance with code generation. All AI-related suggestions have been thoroughly reviewed, and the authors take full responsibility for all writing and all analyses.

The EXPANd trial was made possible by funds from the Swedish Research Council (2016–01965 and 2022-00636), the Parkinson’s Research Foundation, the Swedish Parkinson Foundation as well as the Regional Agreement on Medical Training and Clinical Research (ALF) and the Center for Innovative Medicine (CIMED) - Karolinska Institutet and Region Stockholm. The Research School in Health Care Science and the Strategic Research Area Health Care Science at Karolinska Institute also provided funding.

## Author contributions

E.F. designed the study along with H.J and M.F. M.F., H.J and E.F. were involved in the data collection. M.F., performed the analyses with great help from F.A. with the fMRI analyses. All authors contributed to the interpretation of the results. M.F. drafted the manuscript with input from all authors. All authors read and approved the final manuscript.

