## Supplementary material for "Questioning dual-task assumptions in Parkinson’s disease: heterogeneity, cognitive contributions and adaptative changes"

### SUPPLEMENTARY INFORMATION

#### **Table 1 |** Single and dual task descriptive data

| Outcome | Single task PD | Dual task PD | Single task HC | Dual task HC |
|---|---|---|---|---|
| **Walking task** | | | | |
| Gait speed (m/s) | 1.19 (0.19) | 1.13 (0.23) | 1.39 (0.22) | 1.28 (0.22) |
| Step time variability (ms) | 14.39 (7.06) | 16.53 (10.56) | 11.48 (3.34) | 12.73 (6.44) |
| Reaction time (ms) | 0.97 (0.23) | 0.94 (0.22) | 0.92 (0.27) | 0.95 (0.2) |
| RT variability (ms) | 0.25 (0.22) | 0.23 (0.16) | 0.22 (0.16) | 0.19 (0.1) |
| Step length (m) | 0.65 (0.09) | 0.62 (0.1) | 0.73 (0.07) | 0.69 (0.08) |
| Swing time variability (ms) | 14.28 (6.49) | 16.04 (8.79) | 12.32 (3.65) | 12.45 (7.06) |
| Step time (ms) | 544.5 (40.5) | 548 (55) | 532.5 (56.5) | 545.25 (76.37) |
| Swing time (ms) | 392.5 (34) | 385.5 (42) | 398.75 (23.75) | 399.25 (40) |
| Stance time (ms) | 694.5 (71) | 705.5 (97.5) | 675 (85.62) | 681.75 (110.87) |
| Gait speed variability (m/s) | 0.04 (0.01) | 0.05 (0.02) | 0.04 (0.01) | 0.04 (0.01) |
| Step length variability (m) | 0.02 (0.01) | 0.02 (0.01) | 0.02 (0) | 0.02 (0.01) |
| Stance time variability (ms) | 16.49 (6.61) | 20.24 (12.92) | 12.99 (4.76) | 15.99 (9.61) |
| Swing time asymmetry (ms) | 9 (11) | 10 (10) | 5.5 (8.5) | 8 (8) |
| Step time asymmetry (ms) | 12 (14) | 13 (15) | 8 (10.25) | 10 (14.5) |
| Stance time asymmetry (ms) | 9 (10) | 10 (12) | 5 (6.5) | 8 (8.25) |
| Step length asymmetry (m) | 0.02 (0.02) | 0.02 (0.02) | 0.02 (0.02) | 0.01 (0.02) |
| Step width (m) | 0.08 (0.03) | 0.09 (0.03) | 0.09 (0.03) | 0.09 (0.03) |
| Step width variability (m) | 0.02 (0.01) | 0.02 (0.01) | 0.02 (0.01) | 0.02 (0.01) |
| Accuracy (%) | 100 (6.67) | 96.55 (8.7) | 100 (0) | 100 (5.34) |
| **Scanner task** | | | | |
| RT (ms) | 0.64 (0.12) | 0.72 (0.11) | 0.53 (0.11) | 0.67 (0.12) |
| RT variability (ms) | 0.19 (0.11) | 0.28 (0.1) | 0.14 (0.07) | 0.21 (0.09) |
| Count task (points) | | 8 (6) | | 12 (5.5) |
| Accuracy (%) | 93.75 (14.58) | 77.19 (27.19) | 97.92 (5.21) | 93.75 (14.22) |
| **Timed Up and Go task** | | | | |
| Time (s) | 10.37 (3.05) | 14.41 (5.72) | 8.44 (1.54) | 11.02 (4.28) |

#### **Table 2 |** Dual task model estimates, all values included

| Outcome | Estimate | Test | p | p adjusted |
|---|---|---|---|---|
| **Walking task** | | | | |
| Gait speed m/s | 0.05 (-0.0002, 0.1) | gaussian | 0.05 | - |
| RT | -0.04 (-0.08, 0.0008) | log | 0.05 | - |
| RT variability | 0.003 (-0.2, 0.2) | log | 0.97 | - |
| Step length (m) | 0.01 (-0.003, 0.03) | gaussian | 0.12 | 0.38 |
| Swing time variability (ms) | -0.06 (-0.2, 0.05) | log heterog | 0.29 | 0.52 |
| Step time (ms) | 0.03 (-0.02, 0.07) | gamma | 0.3 | 0.52 |
| Swing time (ms) | -4 (-10, 4) | gaussian | 0.33 | 0.52 |
| Stance time (ms) | -0.02 (-0.05, 0.02) | log | 0.32 | 0.52 |
| Gait speed variability (m/s) | 0.03 (-0.09, 0.1) | log_heterog | 0.63 | 0.84 |
| Swing time asymmetry (ms) | -0.2 (-0.7, 0.3) | sqrt gaussian | 0.48 | 0.7 |
| Step time asymmetry (ms) | -0.6 (-1, -0.03) | sqrt gaussian | 0.04 | 0.32 |
| Stance time asymmetry (ms) | -0.5 (-1, 0.04) | sqrt gaussian | 0.07 | 0.37 |
| Step length asymmetry (m) | 0.02 (0.003, 0.04) | sqrt gaussian | 0.02 | 0.32 |
| Step width (m) | -0.0002 (-0.003, 0.003) | gaussian | 0.9 | 0.9 |
| Step width variability (m) | 0.06 (-0.02, 0.1) | log | 0.15 | 0.4 |
| Step time variability | -0.01 (-0.2, 0.2) | gaussian | 0.88 | 0.9 |
| Step length variability (m) | 0.1 (-0.03, 0.2) | log heterog | 0.12 | 0.38 |
| Stance time variability (ms) | -0.02 (-0.2, 0.1) | log heterog | 0.82 | 0.9 |
| Accuracy (%) | 0.5 (0.4, 0.6) | brunner munzel | 0.84 | - |
| **Scanner task** | | | | |
| RT | 0.09 (0.06, 0.1) | gaussian | 0 | - |
| RT variability | -0.01 (-0.1, 0.1) | log | 0.88 | - |
| Accuracy (%) | 0.7 (0.6, 0.8) | brunner munzel | 0 | - |
| Counting task | 0.3 (0.2, 0.4) | brunner munzel | 0 | - |
| **Timed Up and Go task** | | | | |
| Time (s) | 0.5 (0.4, 0.6) | brunner munzel | 0.68 | 0.84 |

All outcomes analysed with MLM-models except the outcomes where Test = B.M. test i.e., Brunner-Munzel test. For the MLM-models Test indicate the assumed distribution in the model. Log = MLM model based on logged values, assuming gaussian distribution, Log heterog = MLM model based on logged values, assuming gaussian distribution while allowing large heterogeneity between groups and task type. Sqrt gaussian = MLM model based on squared values, assuming gaussian distribution. Estimate = unstandardized b for MLM models. p adjusted = p adjusted with false discovery rate method, only used for secondary values. Group diff. = PD DTE – HC DTE, pp = percent points.

#### **Fig. 1 |** Secondary outcomes presented for the HC group and PD participants with differing cognitive function

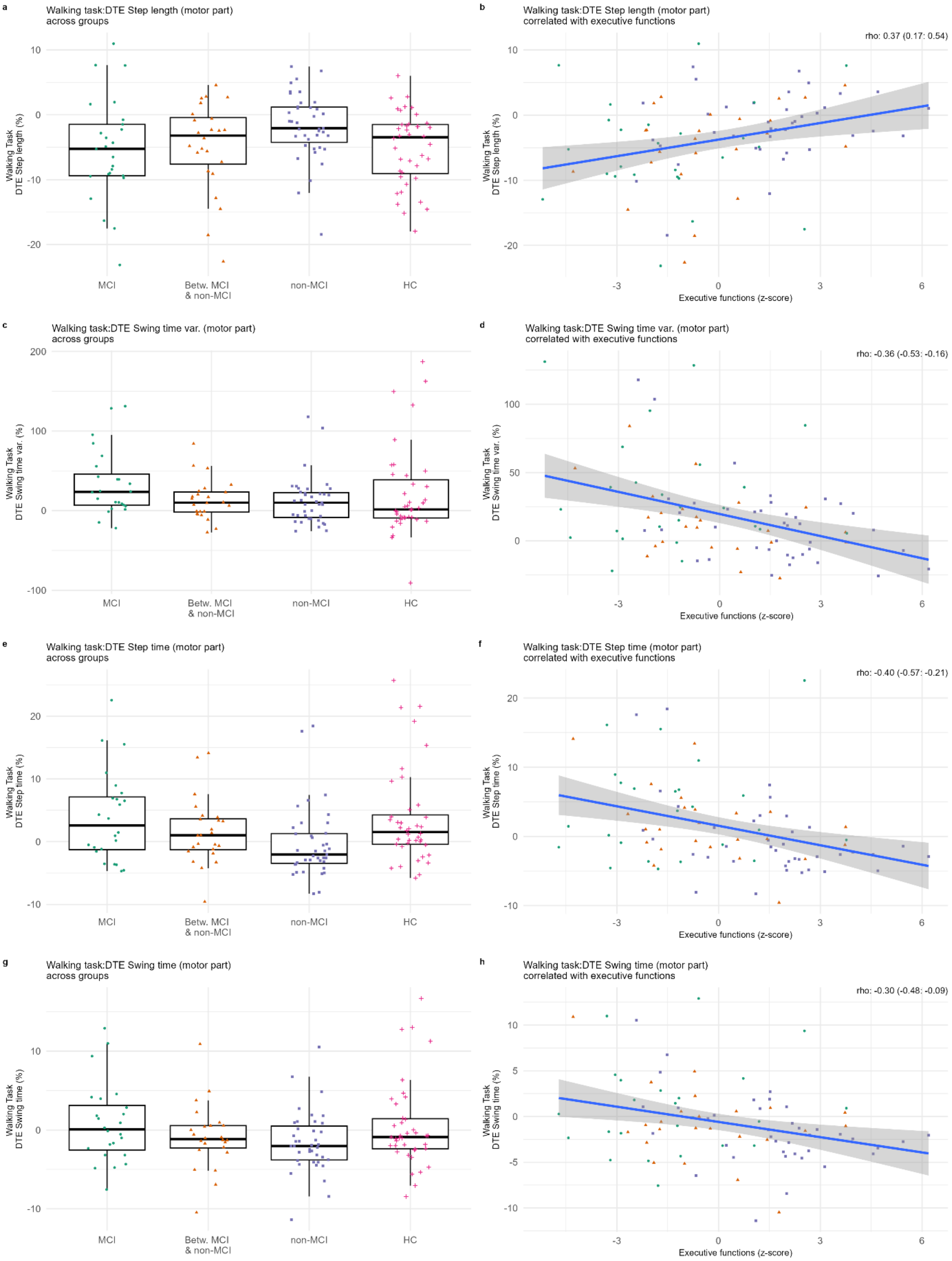

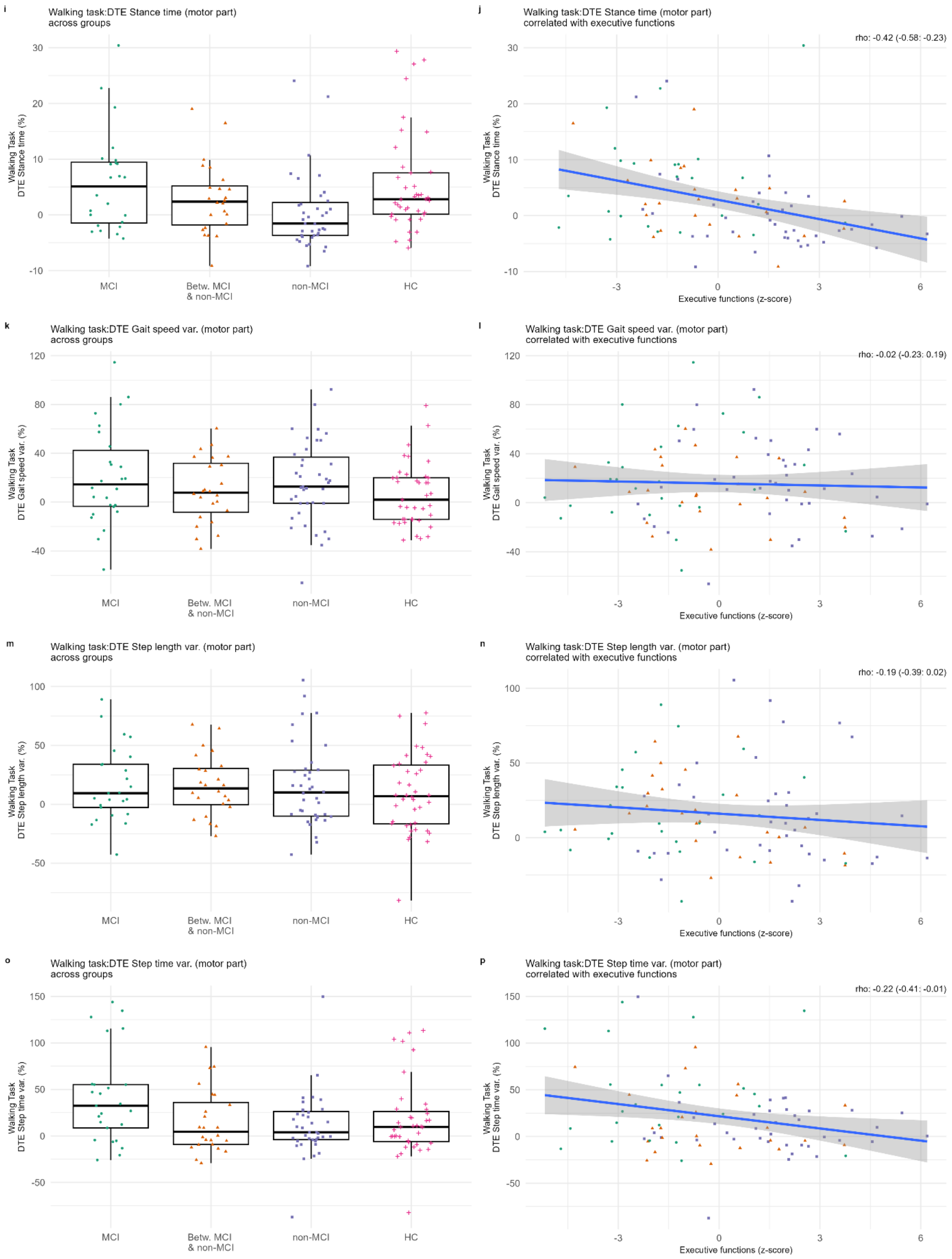

i
Walking task:DTE Stance time (motor part)
across groups
Walking Task
DTE Stance time (%)
30
20
10
0
-10
MCI
Betw. MCI
& non-MCI
non-MCI
HC
j
Walking task:DTE Stance time (motor part)
correlated with executive functions
rho: -0.42 (-0.58: -0.23)
Executive functions (z-score)
-3
0
3
6
k
Walking task:DTE Gait speed var. (motor part)
across groups
Walking Task
DTE Gait speed var. (%)
120
80
40
0
-40
l
Walking task:DTE Gait speed var. (motor part)
correlated with executive functions
rho: -0.02 (-0.23: 0.19)
m
Walking task:DTE Step length var. (motor part)
across groups
Walking Task
DTE Step length var. (%)
100
50
0
-50
n
Walking task:DTE Step length var. (motor part)
correlated with executive functions
rho: -0.19 (-0.39: 0.02)
o
Walking task:DTE Step time var. (motor part)
across groups
Walking Task
DTE Step time var. (%)
150
100
50
0
-50
p
Walking task:DTE Step time var. (motor part)
correlated with executive functions
rho: -0.22 (-0.41: -0.01)

q

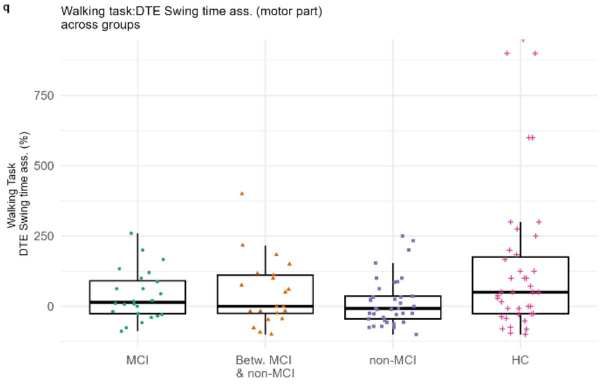
Walking task:DTE Swing time ass. (motor part)
across groups
Walking Task
DTE Swing time ass. (%)
MCI
Betw. MCI
& non-MCI
non-MCI
HC

r

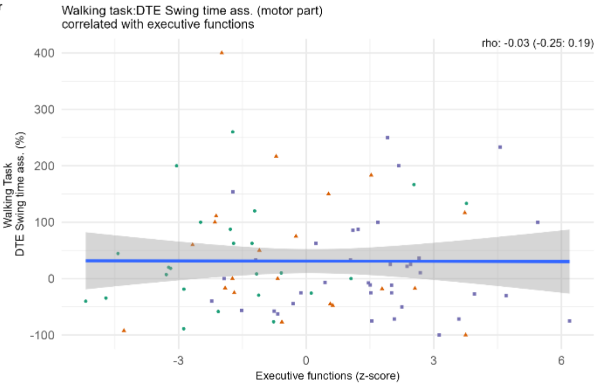
Walking task:DTE Swing time ass. (motor part)
correlated with executive functions
rho: -0.03 (-0.25: 0.19)
Walking Task
DTE Swing time ass. (%)
Executive functions (z-score)

s

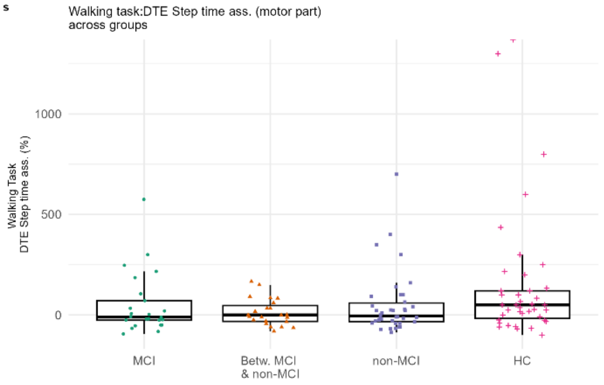
Walking task:DTE Step time ass. (motor part)
across groups
Walking Task
DTE Step time ass. (%)
MCI
Betw. MCI
& non-MCI
non-MCI
HC

t

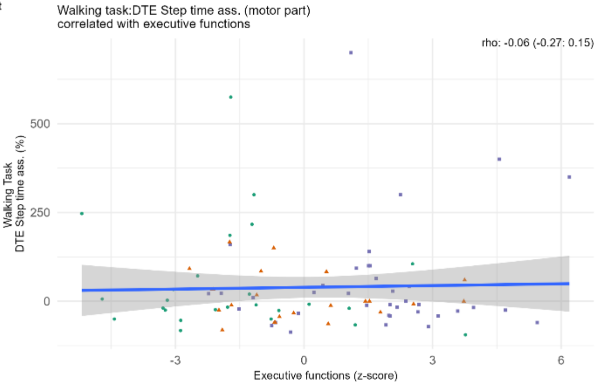
Walking task:DTE Step time ass. (motor part)
correlated with executive functions
rho: -0.06 (-0.27: 0.15)
Walking Task
DTE Step time ass. (%)
Executive functions (z-score)

u

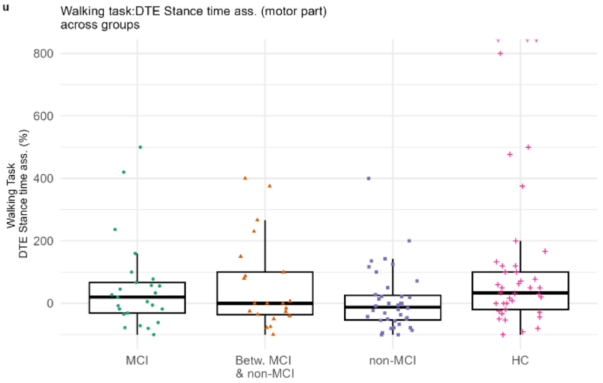
Walking task:DTE Stance time ass. (motor part)
across groups
Walking Task
DTE Stance time ass. (%)
MCI
Betw. MCI
& non-MCI
non-MCI
HC

v

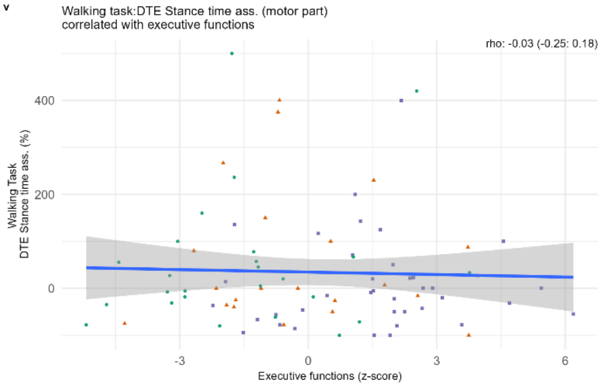
Walking task:DTE Stance time ass. (motor part)
correlated with executive functions
rho: -0.03 (-0.25: 0.18)
Walking Task
DTE Stance time ass. (%)
Executive functions (z-score)

w

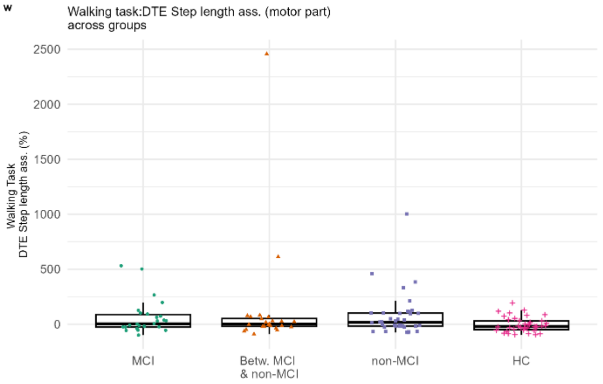
Walking task:DTE Step length ass. (motor part)
across groups
Walking Task
DTE Step length ass. (%)
MCI
Betw. MCI
& non-MCI
non-MCI
HC

x

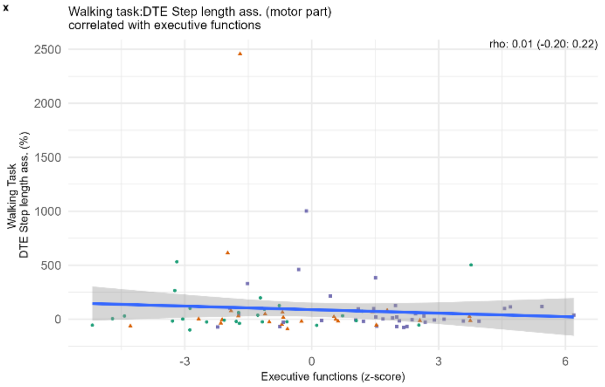
Walking task:DTE Step length ass. (motor part)
correlated with executive functions
rho: 0.01 (-0.20: 0.22)
Walking Task
DTE Step length ass. (%)
Executive functions (z-score)

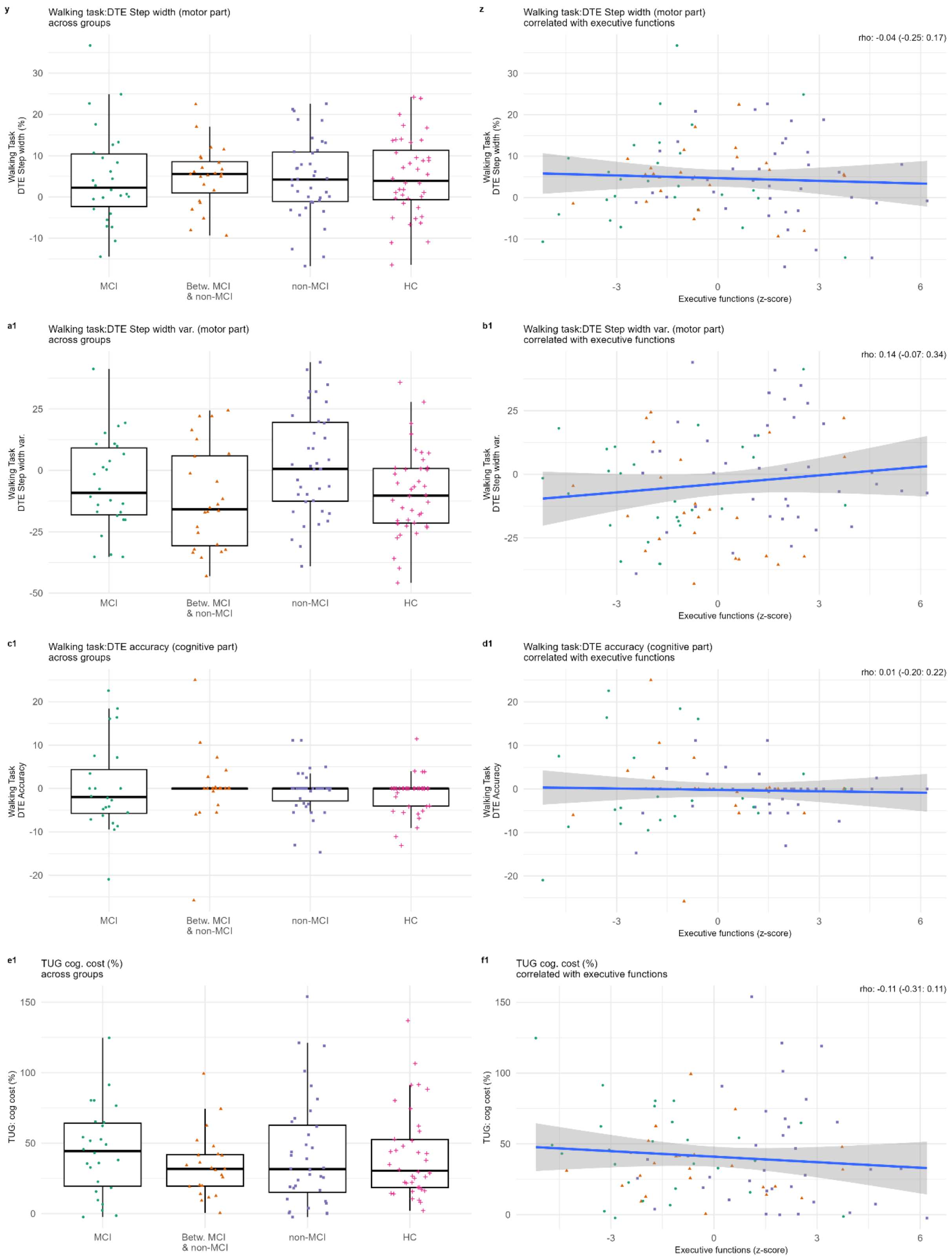
y
Walking task:DTE Step width (motor part)
across groups
Walking Task
DTE Step width (%)
30
20
10
0
-10
MCI
Betw. MCI
& non-MCI
non-MCI
HC
z
Walking task:DTE Step width (motor part)
correlated with executive functions
rho: -0.04 (-0.25: 0.17)
Executive functions (z-score)
-3
0
3
6
a1
Walking task:DTE Step width var. (motor part)
across groups
Walking Task
DTE Step width var.
25
0
-25
-50
b1
Walking task:DTE Step width var. (motor part)
correlated with executive functions
rho: 0.14 (-0.07: 0.34)
c1
Walking task:DTE accuracy (cognitive part)
across groups
Walking Task
DTE Accuracy
20
10
0
-10
-20
d1
Walking task:DTE accuracy (cognitive part)
correlated with executive functions
rho: 0.01 (-0.20: 0.22)
e1
TUG cog. cost (%)
across groups
TUG: cog cost (%)
150
100
50
0
f1
TUG cog. cost (%)
correlated with executive functions
rho: -0.11 (-0.31: 0.11)

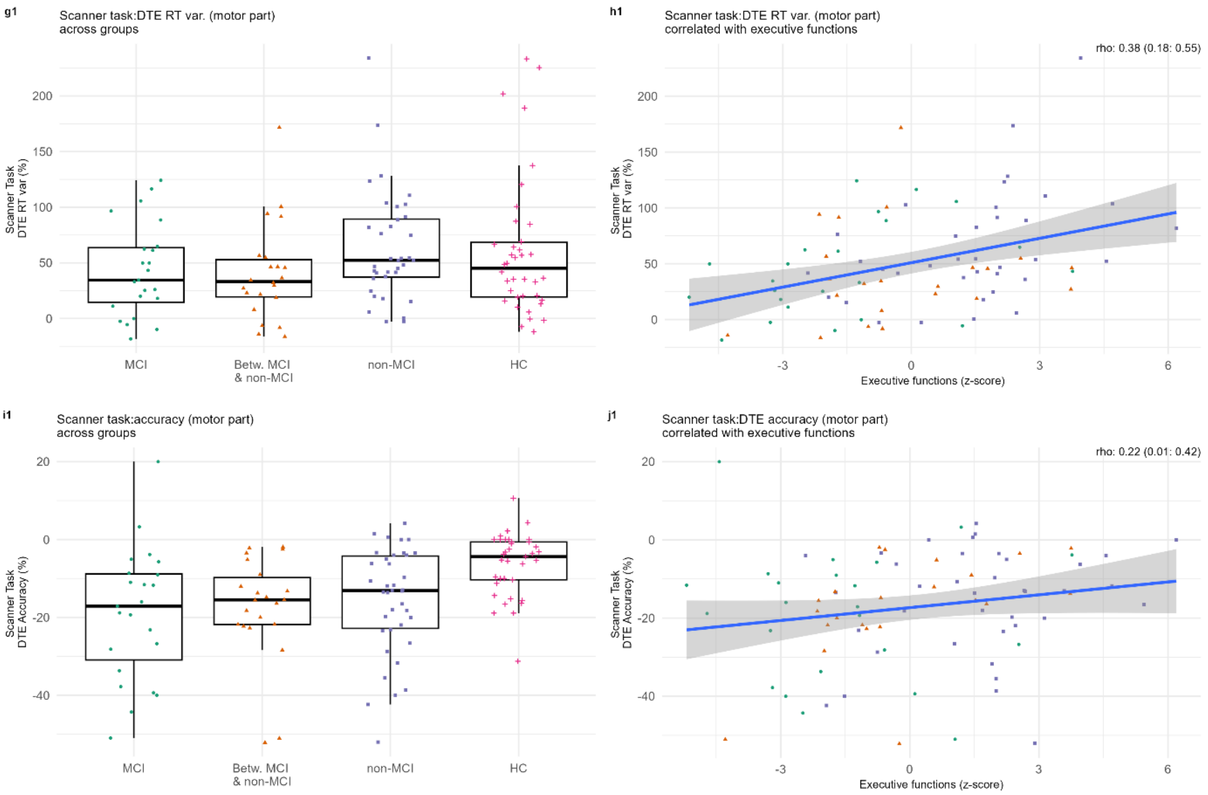

For all boxplots, the PD group is presented classified as with MCI (mild cognitive impairment) or without (non-MCI), or in between those groups (Betw. MCI & non-MCI). HC = healthy control group. All correlation plots are within the PD group. DTE = dual-task effect. rho = Spearman's rang order correlation coefficient.  Var. = variability, RT = reaction time, corr. = correlation. **a-d1** Secondary outcome variables of the Walking task. Executive functions (z-score) = a higher value indicates higher executive functions, **e1-f1** TUG outcomes, **g1**- **j1** Secondary outcome variables of the Scanner task.

#### **Fig. 2 |** Primary outcomes predicted by executive function and task type

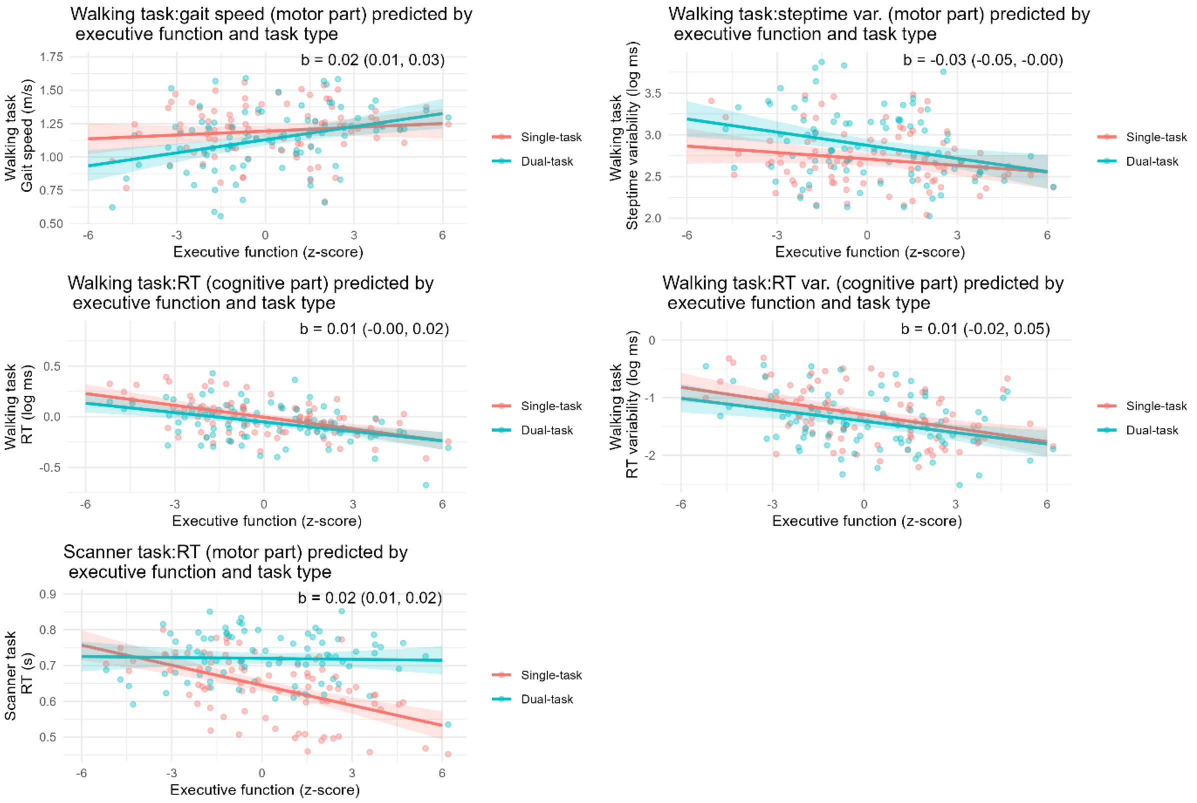

The lines represent predicted values from the multilevel models. Points represent observed data. All models included the predictors executive function, task type (0 = single, 1 = dual) and their interaction. b = the interaction coefficient, with 95% confidence interval, of task and executive function. A higher z-score represents higher executive functions.

#### **Fig. 3 |** Walking task: correlations of DTE RT and possible indicators of gait instability

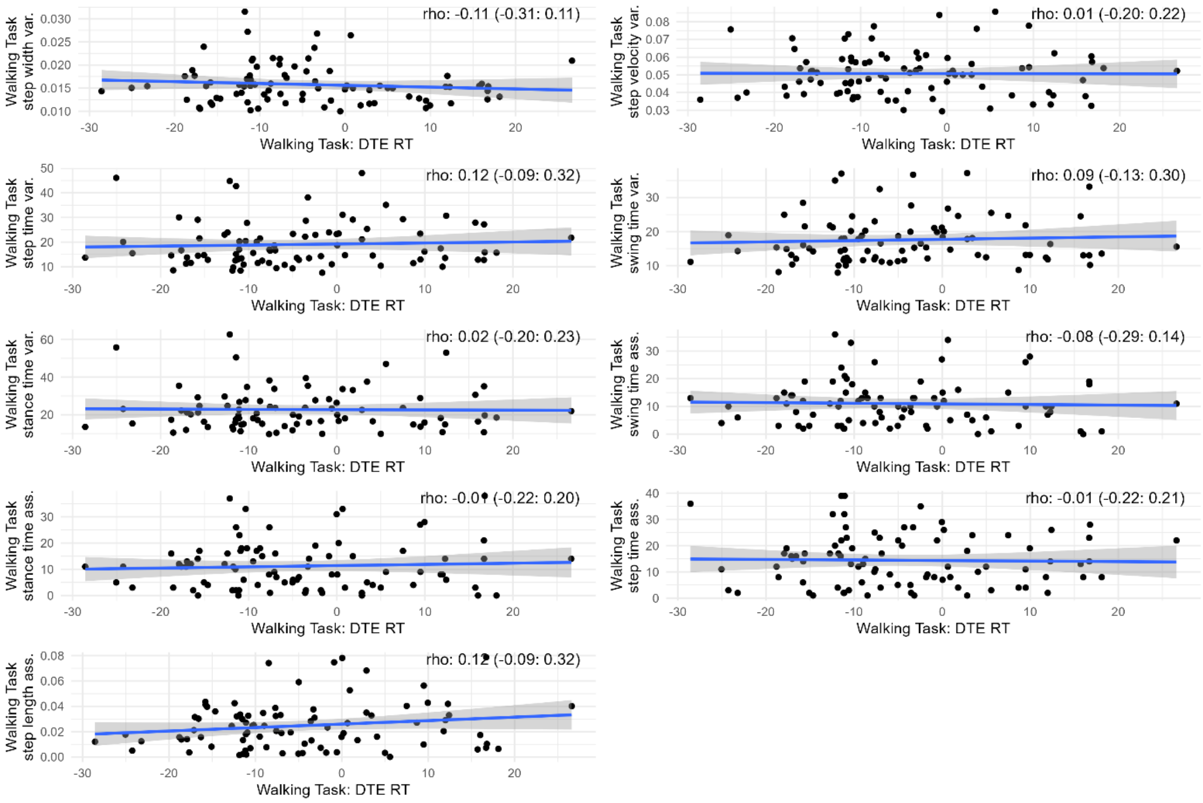

DTE: dual task effect, RT: reaction time, var: variability, ass: assymetry

#### **Table 3 |** Correlations between the tasks: DTE, single and dual outcomes

| | | Walking task | | | | | | | | | Scanner task | | | | TUG | | | Ex. func. |
|---|---|---|---|---|---|---|---|---|---|---|---|---|---|---|---|---|---|---|
| | | Gait speed S | Gait speed D | DTE Gait speed | Step time var. S | Step time var. D | DTE steptime var. | RT var. S | RT var. D | DTE RT var. | Single RT | Dual RT | DTE (%) | Count task | TUG | TUG: Cog | TUG: Cog cost(%) | Ex.func. |
| **Walking task** | WT:Gait speed S | 1 | 0.79 | -0.08 | -0.71 | -0.56 | -0.05 | -0.13 | -0.34 | -0.21 | -0.29 | -0.2 | 0.2 | 0.16 | -0.65 | -0.41 | 0.09 | 0.12 |
| | Gait speed D | 0.79 | 1 | 0.47 | -0.62 | -0.77 | -0.4 | -0.32 | -0.48 | -0.22 | -0.32 | -0.18 | 0.25 | 0.2 | -0.64 | -0.41 | -0.02 | 0.34 |
| | DTE Gait speed | -0.08 | 0.47 | 1 | -0.03 | -0.47 | -0.69 | -0.39 | -0.35 | -0.06 | -0.12 | -0.06 | 0.11 | 0.08 | -0.11 | -0.06 | -0.15 | 0.41 |
| | Step time var. S | -0.71 | -0.62 | -0.03 | 1 | 0.67 | -0.04 | 0.13 | 0.31 | 0.3 | 0.31 | 0.14 | -0.28 | -0.26 | 0.65 | 0.37 | 0 | -0.19 |
| | Step time var. D | -0.56 | -0.77 | -0.47 | 0.67 | 1 | 0.58 | 0.27 | 0.46 | 0.19 | 0.31 | 0.14 | -0.27 | -0.18 | 0.45 | 0.38 | 0.06 | -0.26 |
| | DTE steptime var. | -0.05 | -0.4 | -0.69 | -0.04 | 0.58 | 1 | 0.37 | 0.33 | 0.04 | 0.2 | 0.09 | -0.18 | -0.05 | 0.12 | 0.14 | 0.16 | -0.26 |
| | RT var. S | -0.13 | -0.32 | -0.39 | 0.13 | 0.27 | 0.37 | 1 | 0.46 | -0.14 | 0.39 | 0.16 | -0.38 | -0.03 | 0.25 | 0.14 | 0.02 | -0.53 |
| | RT var. D | -0.34 | -0.48 | -0.35 | 0.31 | 0.46 | 0.33 | 0.46 | 1 | 0.39 | 0.29 | 0.2 | -0.19 | -0.09 | 0.28 | 0.32 | 0.15 | -0.34 |
| | DTE RT var. | -0.21 | -0.22 | -0.06 | 0.3 | 0.19 | 0.04 | -0.14 | 0.39 | 1 | -0.03 | -0.02 | 0.02 | -0.2 | 0.34 | 0.11 | -0.1 | 0.02 |
| **Scanner task** | Single RT | -0.29 | -0.32 | -0.12 | 0.31 | 0.31 | 0.2 | 0.39 | 0.29 | -0.03 | 1 | 0.69 | -0.72 | -0.28 | 0.43 | 0.29 | -0.02 | -0.53 |
| | Dual RT | -0.2 | -0.18 | -0.06 | 0.14 | 0.14 | 0.09 | 0.16 | 0.2 | -0.02 | 0.69 | 1 | 0.01 | -0.19 | 0.26 | 0.19 | 0 | -0.03 |
| | Dte cost (%) | 0.2 | 0.25 | 0.11 | -0.28 | -0.27 | -0.18 | -0.38 | -0.19 | 0.02 | -0.72 | 0.01 | 1 | 0.2 | -0.33 | -0.22 | 0.03 | 0.61 |
| | Count task | 0.16 | 0.2 | 0.08 | -0.26 | -0.18 | -0.05 | -0.03 | -0.09 | -0.2 | -0.28 | -0.19 | 0.2 | 1 | -0.33 | -0.26 | -0.23 | 0.26 |
| **TUG** | TUG | -0.65 | -0.64 | -0.11 | 0.65 | 0.45 | 0.12 | 0.25 | 0.28 | 0.34 | 0.43 | 0.26 | -0.33 | -0.33 | 1 | 0.58 | -0.02 | -0.25 |
| | TUG:Cog | -0.41 | -0.41 | -0.06 | 0.37 | 0.38 | 0.14 | 0.14 | 0.32 | 0.11 | 0.29 | 0.19 | -0.22 | -0.26 | 0.58 | 1 | 0.64 | -0.2 |
| | TUG:Cog cost(%) | 0.09 | -0.02 | -0.15 | 0 | 0.06 | 0.16 | 0.02 | 0.15 | -0.1 | -0.02 | 0 | 0.03 | -0.23 | -0.02 | 0.64 | 1 | -0.1 |
| **Ex. func.** | Ex.func. | 0.12 | 0.34 | 0.41 | -0.19 | -0.26 | -0.26 | -0.53 | -0.34 | 0.02 | -0.53 | -0.03 | 0.61 | 0.26 | -0.25 | -0.2 | -0.1 | 1 |

S: single task, D: dual task, TUG:timed up and go, Ex. func: executive functions, Cog:cognitive part, DTE: dual task effect, RT: reaction time, var: variability,

**Fig.4 |** Correlation of putamen activity and Scanner DTE RT within the HC group

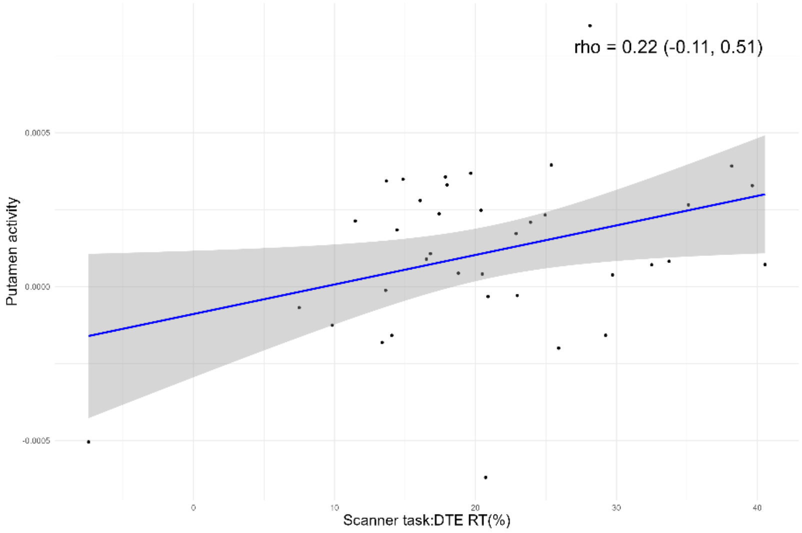

DTE = dual-task effect. rho = Spearman's rang order correlation coefficient. RT = reaction time.
